# Development and initial evaluation of a conversational agent for Alzheimer’s disease

**DOI:** 10.1101/2024.09.04.24312955

**Authors:** Natalia Castano-Villegas, Isabella Llano, Maria Camila Villa, Julian Martinez, Jose Zea, Tatiana Urrea, Alejandra Maria Bañol, Carlos Bohorquez, Nelson Martinez

## Abstract

**Background:** Conversational Agents have attracted attention for personal and professional use. Their specialisation in the medical field is being explored. Conversational Agents (CA) have accomplished passing-level performance in medical school examinations and shown empathy when responding to patient questions. Alzheimer’s disease is characterized by the progression of cognitive and somatic decline. As the leading cause of dementia in the elderly, it is the subject of continuous investigations, which result in a constant stream of new information. Physicians are expected to keep up with the latest clinical guidelines; however, they aren’t always able to do so due to the large amount of information and their busy schedules.

**Objective:** We designed a conversational agent intended for general physicians as a tool for their everyday practice to offer validated responses to clinical queries associated with Alzheimer’s Disease based on the best available evidence.

**Methodology:** The conversational agent uses GPT-4o and has been instructed to respond based on 17 updated national and international clinical practice guidelines about Dementia and Alzheimer’s Disease. To approach the CA’s performance and accuracy, it was tested using three validated knowledge scales. In terms of evaluating the content of each of the assistant’s answers, a human evaluation was conducted in which 7 people evaluated the clinical understanding, retrieval, clinical reasoning, completeness, and usefulness of the CA’s output.

**Results:** The agent obtained near-perfect performance in all three scales. It achieved a sensitivity of 100% for all three scales and a specificity of 75% in the less specific model. However, when modifying the input given to the assistant (prompting), specificity reached 100%, with a Cohen’s kappa of 1 in all tests. The human evaluation determined that the CA’s output showed comprehension of the clinical question and completeness in its answers. However, reference retrieval and perceived helpfulness of the CA reply was not optimal.

**Conclusions:** This study demonstrates the potential of the agent and of specialised LLMs in the medical field as a tool for up-to-date clinical information, particularly when medical knowledge is becoming increasingly vast and ever-changing. Validations with health care experts and actual clinical use of the assistant by its target audience is an ongoing part of this project that will allow for more robust and applicable results, including evaluating potential harm.

## INTRODUCTION

Natural Language Processing (NLP) is an area of computer science that focuses on converting written and spoken natural human languages into structured data [1]. This area has led to the development of large language models (LLM) like the Generative Pre-trained Transformer 3.5 (GPT-3.5), 4 (GPT-4), and 4o (GPT-4o) used in the commonly known chatbot ChatGPT, as well as the Pathways Language Model 2 (PaLM 2) and Large Language Model Meta AI 2 (LLaMA 2). These are trained on datasets containing hundreds of billions of sources from articles, books and the internet and have revolutionised the field of NLP, attracting attention for personal and professional use, specifically in the form of conversational agents (CAs), which are programs designed to engage in human-like conversations with users [2]. In the medical field, ChatGPT has demonstrated passing-level performance in medical school examinations and proven capable of empathy when responding to patient questions [3].

Alzheimer’s disease (AD) refers to the specific onset and progression of cognitive and somatic decline; it is the leading cause of dementia in individuals over 65 years of age [4]. The progressive accumulation of extracellular amyloid beta plaques and intracellular neurofibrillary tangles characterises AD. As the disease progresses and cognitive impairment worsens, patients experience severe behavioural symptoms that result in the need for round-the-clock care. The early detection of Alzheimer’s disease symptoms allows patients to be part of the development of care plans before they deteriorate, seek early interventions and apply lifestyle changes that can help maintain quality of life as long as possible [4]. Research is being continuously conducted to find effective treatments for AD, which currently has no cure. This implies a constant stream of new information regarding diagnosis, treatment, new medication, and chronic management, among others, requiring clinicians to stay updated.

The development of CAs for specifi” tas’s and fields in medicine has been explored, including, but not limited to, radiology [5], oncology [6], and ophthalmology [3]. However, to the best of our knowledge, no CAs have focused on Alzheimer’s disease. AD presents a particular interest as the fifth leading cause of death globally, with a growing prevalence among adults in their late working or retirement years of 3-4%. AD can often be challenging to diagnose as cognitive decline due to ageing, and other types of dementia can pose similar features [7]. The importance of risk detection is emphasised by studies that state that treatments are more effective when administered before the emergence of dementia [8]. Primary care clinicians are often the patient’s first contact in the healthcare system. They are, therefore, critical in determining the nature of the approach needed to adequately guide the subject and their family through the next steps of diagnosis and care. However, according to the 2020 Alzheimer’s Association Primary Care Physician Dementia Care Training Survey [9], 53% of clinicians recognise they only keep up with new developments “very little” or “not at all,” despite 93% acknowledging it was their duty to stay informed. AI conversational agents can be adapted to aid students, trainees, and professionals from healthcare-related fields in staying on top of current developments and guidelines. Consequently, we aimed to develop a CA that can offer instantaneous, accurate, and validated responses to clinical queries associated with AD from a comprehensive set of clinical practice guidelines for dementias, specifically Alzheimer’s disease, intended for general physicians, as a tool for their everyday consultation.

## METHODOLOGY

### Conversational Agent Development

The Dementia-Alzheimer’s Conversational Agent (DACA) was developed and programmed based on the information found within 17 Clinical Practice Guidelines, including 9 for dementia in general and 7 for Alzheimer’s Disease. The bibliography includes guidelines in English (11 references) and Spanish (6 references). It contains information on diagnosis, management, treatment, medications, disease prevalence, prognosis, biomarkers, risk factors, epidemiology, patient support, primary care, psychosocial interventions and palliative care (Table 1.). With the support of the team’s experts in neurology and AD, guidelines were chosen based on their relevance in clinical practice. The literature used was mainly provided by the educational program coordinated by national Alzheimer’s Disease experts. This program is designed to train general physicians and raise awareness of Alzheimer’s disease and is executed by the pharmaceutical Knight in Colombian insurance companies (EPS).

**Table 1:**
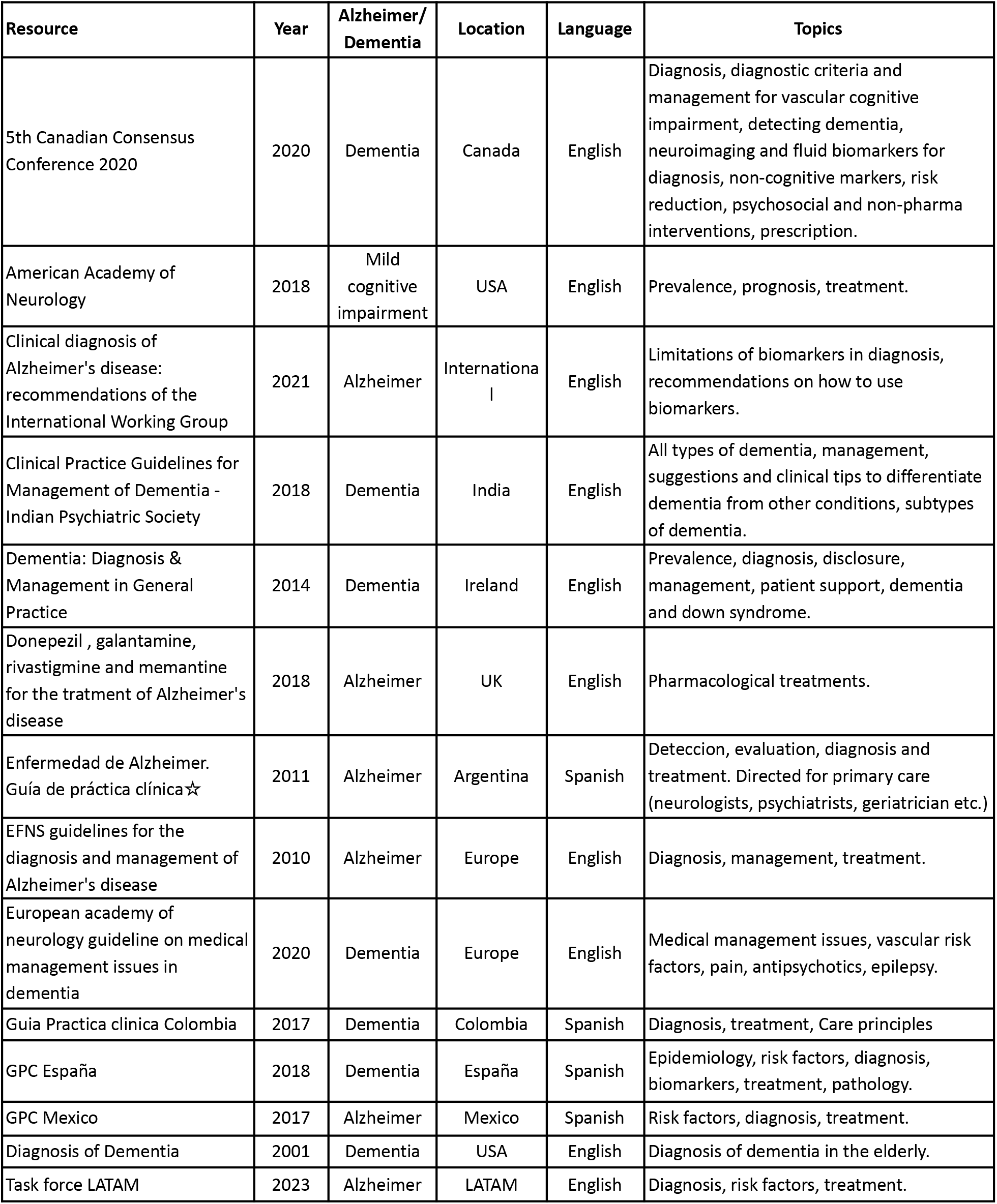
Resources included in the knowledge base for the DACA.

| Resource | Year | Alzheimer/<br>Dementia | Location | Language | Topics |
| --- | --- | --- | --- | --- | --- |
| 5th Canadian Consensus Conference 2020 | 2020 | Dementia | Canada | English | Diagnosis, diagnostic criteria and management for vascular cognitive impairment, detecting dementia, neuroimaging and fluid biomarkers for diagnosis, non-cognitive markers, risk reduction, psychosocial and non-pharma interventions, prescription. |
| American Academy of Neurology | 2018 | Mild cognitive impairment | USA | English | Prevalence, prognosis, treatment. |
| Clinical diagnosis of Alzheimer's disease: recommendations of the International Working Group | 2021 | Alzheimer | International | English | Limitations of biomarkers in diagnosis, recommendations on how to use biomarkers. |
| Clinical Practice Guidelines for Management of Dementia - Indian Psychiatric Society | 2018 | Dementia | India | English | All types of dementia, management, suggestions and clinical tips to differentiate dementia from other conditions, subtypes of dementia. |
| Dementia: Diagnosis & Management in General Practice | 2014 | Dementia | Ireland | English | Prevalence, diagnosis, disclosure, management, patient support, dementia and down syndrome. |
| Donepezil , galantamine, rivastigmine and memantine for the tratment of Alzheimer's disease | 2018 | Alzheimer | UK | English | Pharmacological treatments. |
| Enfermedad de Alzheimer. Guía de práctica clínica☆ | 2011 | Alzheimer | Argentina | Spanish | Deteccion, evaluation, diagnosis and treatment. Directed for primary care (neurologists, psychiatrists, geriatrician etc.) |
| EFNS guidelines for the diagnosis and management of Alzheimer's disease | 2010 | Alzheimer | Europe | English | Diagnosis, management, treatment. |
| European academy of neurology guideline on medical management issues in dementia | 2020 | Dementia | Europe | English | Medical management issues, vascular risk factors, pain, antipsychotics, epilepsy. |
| Guia Practica clinica Colombia | 2017 | Dementia | Colombia | Spanish | Diagnosis, treatment, Care principles |
| GPC España | 2018 | Dementia | España | Spanish | Epidemiology, risk factors, diagnosis, biomarkers, treatment, pathology. |
| GPC Mexico | 2017 | Alzheimer | Mexico | Spanish | Risk factors, diagnosis, treatment. |
| Diagnosis of Dementia | 2001 | Dementia | USA | English | Diagnosis of dementia in the elderly. |
| Task force LATAM | 2023 | Alzheimer | LATAM | English | Diagnosis, risk factors, treatment. |

**Table 2.** Knowledge scale scores to both experiments. Present the assistant with only the statement and provide instructions to indicate whether the statement is True or False.

| Scale | Statements only |  |  | Prompt: "Is this statement True or False?" |  |  | Number of questions (n) | Average response time (seconds) |
| --- | --- | --- | --- | --- | --- | --- | --- | --- |
| | # correct answers (n) | Score | Cohen's Kappa ($\kappa$) | # correct answers (n) | Score | Cohen's Kappa ($\kappa$) | | |
| DKAS | 27 | 100.00% | 1.00 | 27 | 100.00% | 1.00 | 27 | 5.82 |
| UJA ACS | 21 | 91.30% | 0.80 | 23 | 100.00% | 1.00 | 23 | 6.42 |
| ADKS | 28 | 93.33% | 0.86 | 30 | 100.00% | 1.00 | 30 | 4.74 |

### Artificial intelligence methods

#### LLM Architectures

The LLM architectures consisted mainly of the Generative Pre-trained Transformer (GPT) family, including 4o (GPT-4o). GPT refers to a pre-trained language model that can generate human-like text and content and respond to questions conversationally. This model can be further developed to execute specific tasks by embedding instructions into the model so the user can obtain coherent, immediate responses without providing any information apart from the actual clinical query.

To this end, our CA was instructed to:

1. Generate predetermined responses to specific queries
2. Answer only queries associated with Alzheimer’s and dementia; it cannot be used generically like ChatGPT.
3. Use technical terms to improve comprehension.
4. Use an adequate writing structure, using main titles, subtitles, bold and capital letters
5. Respond to all queries in Spanish
6. Use more than one reference to provide answers
7. Add citations for papers and guidelines used for answering

GPT-3.5, a different version of the same LLM, was also explored and proved to have a shorter response time (<40 seconds). However, the more complex architecture of GPT-4o made it more potent, with better retrieval (see the section below), more rigorous with its answers, and comparable response time. Additionally, we experimented with assistant complexity, using one assistant and an agent architecture, where multiple assistants answer the same question to come to a final answer.

#### Sources

There were three ways in which the system replied. Initially, the assistant answered with the information included in its initial pre-training without any additional specific literature. Responses using this data were simple and general and judged irrelevant in the medical field. Then, more than 40 Papers and Clinical Guidelines on different topics for AD and dementia were given to the model as sources of information and converted to .txt format. Still, this approach evidenced that the models functioned better with shorter, more specific references instead of a wide selection of literature. The third information source we experimented with was made using PubMed’s search engine with a similar conclusion. Based on this, the authors decided to instruct the Assistant with seventeen updated and specific Clinical Guidelines selected based on date, author and expert suggestions.

#### Retrieval Augmented Generation

The technique of CAs to search and find information in specific documents to answer user queries is called Retrieval Augmented Generation. This method combines retrieval-based and generation-based methods to enhance the accuracy and relevance of generated content. By first retrieving pertinent information from a large corpus and then using a generative model to produce coherent text, RAG ensures contextually appropriate and highly accurate responses, making it ideal for applications requiring precision and relevance.

To include information presented as graphs or tables inside clinical guidelines, we transformed the information with Markdown Sintaxis and saved it as a text file (.txt).

### Evaluation

#### Preliminary Assistant Evaluation

Our team’s medical experts and engineers evaluated the responses of all combinations, considering response completeness and correctness, time to respond, response length, and appropriate referencing. Based on this, one model was chosen for further evaluation.

#### Knowledge scale evaluation

Three peer-reviewed validated knowledge scales for dementia evaluation were used to test the conversational agent against a standard. All the scales were open-sourced.

1. The Dementia Knowledge Assessment Scale (DKAS) This scale was developed and published in 2015. It evaluated test-retest reliability, internal consistency, preliminary construct, concurrent, and factorial validity. It comprises 27 questions and is available in multiple languages, including Spanish. This scale is intended to assess knowledge deficiency in people who provide care and treatment to dementia patients [10]. The DKAS was tested on volunteers from a dementia-related course (nurses, professional care workers, family caregivers, physicians, students), medical students in a residential aged care facility and Australian health workers (hospital staff, general practitioners, residents). Cohorts with higher dementia knowledge and experience scored between 59% and 75%, while individuals with less scored around 53% in the DKAS [10]. It has been further validated in an international cohort, revealing good reliability and internal consistency [11]. Version 2 of the DKAS was developed to assess knowledge in families and staff [12]. Both versions are available online and free of charge. We obtained explicit permission from the author to use the scale.
2. The UJA Alzheimer’s Care Scale (UJA ACS) This is a 23-item scale developed in 2019 to test nurses’ and caregivers’ knowledge about caring for dementia patients. The UJA ACS was tested among nursing staff (registered nurses, assistant nurses, and eldercare workers) and nursing students, who obtained average scores of 75% and 67%, respectively [13]. It was evaluated for content and psychometric properties, showing a proper ability to distinguish between professionals with low and high knowledge and appropriate test-retest correlation [13].
3. The Alzheimer’s Disease Knowledge Scale (ADKS) This scale, developed in 2009, includes 30 items and was obtained from one of the authors, Dr. Brain Carpenter. It evaluates knowledge about risk factors, assessment and diagnosis, symptoms, disease course, life impact, caregiving, treatment, and management. The ADKS was tested in students, older adults, senior centre staff, dementia caregivers and dementia professionals, who obtained, on average, 67%, 80%, 67%, 76% and 91%, respectively [16]. The scale has been shown to have appropriate reliability and validity and is designed for assessing Alzheimer’s knowledge in laypeople, patients, caregivers, and professionals [14].

All three scales include true-or-false statements. In this sense, we determined that agreement between the two methods would be evaluated as total agreement (100%) or complete disagreement (0%).

The assistant was tested by entering the scales’ statements at two different times: one without further instruction and two, writing the instruction, “Answer true or false, according to the following statements”. This rephrasing is known as Prompting. The time to initiate a response within the three scales was measured for each statement and the approaches mentioned. All statements were entered individually and timed from the moment the question was asked until the beginning of the answer. Agreement between scales standard answers and our CA was measured with Cohen’s kappa. Time was measured in seconds. A complete display of knowledge scale statements, CA responses, and standard answers can be provided upon request.

#### Human Evaluation

Seven members of our research team who were not involved in the development of the CA were asked to assess its answers based on clinical reasoning, language and response appropriateness using the following questionnaire, in which each response has a number of points to rate the CA’s answers:

1. Does the answer evidence the conversational agent’s comprehension of the clinical question?
  a. There is no clinical comprehension (1)
  b. There is some mistake in the clinical comprehension (2)
  c. The assistant had perfect clinical comprehension (3)
2. Does the answer demonstrate an adequate ability to retrieve relevant information from bibliographical sources?
  a. No, the information retrieved isn’t relevant to the question asked (1)
  b. The information retrieved could have been more relevant to the question asked (2)
  c. Yes, the information retrieved is entirely relevant to the question asked (3)
3. Does the answer evidence an ability for clinical reasoning?
  a. The question did not require clinical reasoning (1)
  b. No, the assistant did not show any ability for clinical reasoning (2)
  c. Yes, the assistant shows an ability for clinical rationale (3)
4. Does the answer provided respond to the question entirely?
  a. No, the answer provided does not respond to question (1)
  b. The answer provided responds partially to the question (2)
  c. Yes, the answer provided responds the question (3)
5. Was the conversational assistant’s helpful answer for you?
  a. Not at all helpful (1)
  b. Somewhat helpful (2)
  c. Very helpful (3)

The questions were randomly assigned to each evaluator, using a random sequence generator in Python 3.0, in sets of three answers per researcher, without repetition.

An average score was calculated for each question using their final punctuation. We compared the scores for CA answers with the straightforward approach (just providing the statements) and the prompting approach (giving the CA a previous instruction) to evaluate any modifications in response specificity. Table 3. describes the results.

**Table 3.**
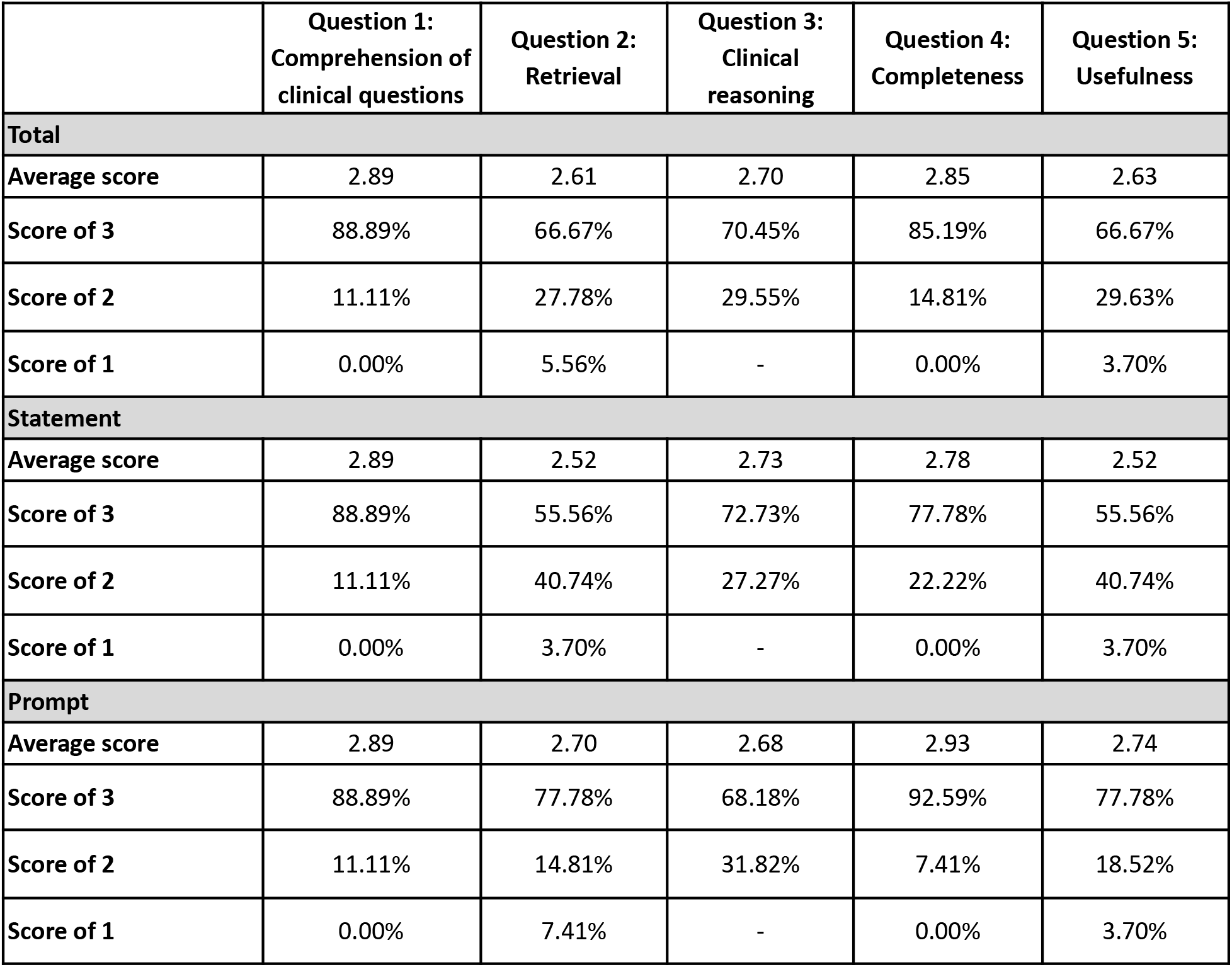
Human evaluation results. Average scores and frequency of perfect score (3) for each question. Results are presented for all evaluated questions and for both approaches: statements only and using the prompt.

## RESULTS

### Conversational Agent Characteristics

The conversational agent chosen used GPT-4o as the LLM architecture, had clinical guidelines as the information source, and was based on one agent instead of multiple ones. This combination provided the best answers regarding quality and retrieval, compared to GPT-3.5, even though it had a shorter response onset time (<40 seconds). Using multiple agents improved response quality but drastically increased the response onset time (over 40 seconds). The experimentation on databases revealed that the assistant had poor performance when loaded with too much information; using PubMed library or other extensive databases generated noise and did not improve the quality of the model’s answers. However, the use of GPT-4o with its baseline knowledge (a wide range of online content from websites to Wikipedia, fiction, non-fiction books and scientific papers) resulted in simple and informal answers that were not adequate for the purpose of the CA.

### Knowledge scale evaluation

The CA performed well on the three knowledge scales when presented with the straightforward approach (see methods), obtaining a response concordance of 100% with Cohen’s κ of 1 in the DKAS, 91.3% (Cohen’s κ = 0.80) in the UJA ACS and 95% (Cohen’s κ = 0.86) in the ADKS (Table 2.). In the DKAS, the assistant incorrectly classified two statements as positive. The assistant’s responses are presented in English for the sake of this paper, original responses in Spanish are available upon reasonable request :

1. **DKAS statement**: “Provide a normal diet while assessing the causes of dysphagia.” **Conversational Assistant (DACA):** Providing a normal diet is crucial while the subjacent causes of dysphagia are being evaluated. It is fundamental to involve a specialist in the evaluation and advice regarding deglution and nutrition in persons with dementia to guarantee an integral approach. **Standardized answer:** False
2. **DKAS statement:** “Use of a nasogastric tube or percutaneous gastrostomy in the patient with advanced dementia as a regular feeding route if dysphagia.”

### Conversational Assistant (DACA)

The use of a nasogastric tube or percutaneous gastrostomy should not be employed as a regular feeding route in patients with advanced dementia except in justified cases when there is a real probability of improvement or maintenance of the patients condition. Always prioritize patient comfort, especially in terminal stages

**Standardized answe**r: False

In the ADKS, the assistant answered two questions wrong, labelling them as True or responding ambiguously when they were False.

1. **ADKS statement:** “When people with AD begin to have difficulty taking care of themselves, caregivers should take over right away.”

### Conversational Assistant (DACA)

\### Assistance for self-caring in Alzheimer

When people with Alzheimer’s Disease (AD) start having difficulties taking care of themselves, caregivers should take over right away to assist.

**Reasons for Early Intervention**

1. **Security**:

-Prevent accidents and injuries.

-Monitoring the administration of medication

2. **General Wellbeing**

-Ensure the patient receives adequate nutrition, hygiene and medical attention

3. **Stress Reduction**

-Alleviate patient anxiety associated with their difficulties in carrying out daily activities.

**Strategies for caregivers**

-**Evaluate Needs**: Identify specific areas where the patient needs help.

-**Establish routines**: Create daily routines that provide structure and stability

-**Emotional Support**: Offer emotional support and understanding to improve the patient’s quality of life.)

**Standardized answer**: False

2. **ADKS statement:** “Most people with AD live in nursing homes.”

### Conversational Assistant (DACA)

###Residence in nursing homes for people with Alzheimer’s

Most people with advanced Alzheimer’s disease (AD) eventually need to live in nursing homes due to the progression of the disease and the need for intensive care.

**Statistics**

-In USA and Europe, between half and two-thirds of nursing home residents have dementia

-The physical and emotional burden on family caregivers is often a deciding factor for institutionalization

**Factors for institutionalization**

1. **Extensive care needs**:

-Including nighttime care and management of incontinence

2. **Physical and mental deterioration**:

-Both the caregiver and the patient

3. **Lack of support**:

-Insufficient formal and informal resources)

**Standardized answer**: False

All of the mistakes made by the CA classified false statements as positive. This affects the model’s specificity, dropping to 75% for the UJA ACS and 83.3% for the ADKS, while sensibility remains consistently at 100%. On the other hand, with the prompting approach (see methods), all four mistakes were corrected, and the assistant achieved 100% performance with 100% sensibility and specificity and a concordance of 100% for all responses. Cohen’s kappa was 1 on all three scales. Table 2 also shows the average response time for each scale, which was approximately 5 to 7 seconds.

### Human Evaluation

Nine questions from each scale (27 total) were evaluated based on five categories applied to both the straightforward and the prompting approaches. In general, our CA obtained scores over 2.5/3 on all questions (Table 3). Question 1 (Q1) about the assistant’s comprehension of clinical questions received the highest punctuation (2.9), meaning that almost 90% of the time, it showed an appropriate understanding of the question. The following punctuation was for Q4 (2.85), in which the answer provided by the CA was judged as “complete” 85% of the time. Q3, regarding whether or not the CA’s answer showed to have made adequate clinical reasoning to respond, was next with an average of 2.7 and 70% success. Usefulness and relevant reference retrieval were the lowest, with 67% each and an average score of 2.6. Retrieval, completeness and usefulness (Q2, Q4, Q5) were perceived to improve when the prompting approach was used, while clinical understanding (Q1) remained the same. However, clinical reasoning (Q3) was perceived to decrease its performance (from an average score of 2.73 with the straightforward approach to 2.68 with the prompting approach).

## DISCUSSION

The DACA demonstrates adequate knowledge and understanding of Alzheimer’s disease and Dementia, obtaining over 90% on the three knowledge scales used as the standard of reference. It consistently agreed better with the standard answers than the highest scores described for tier validation in their targeted human population. Respectively, for the DKAS, the UJA ACS and the ADKS, the score improvement was 35%, 16.3% and 2%, which proves the DACA’s ability to answer clinical questions of dementia and Alzheimer’s disease accurately. It also reflects a good retrieval ability to select information to answer the questions from its knowledge base, constituted of 17 national and international updated clinical guidelines.

Of the 80 statements presented, DACA incorrectly classified four answers (5%) when used with the straightforward approach (statements given without further instruction), as listed in the Knowledge Scale Evaluation in the Results section. This meant that the operational characteristics of the CAl, such as sensitivity and specificity, were 100% and 83%, respectively, and the Positive Predictive value was 90%.

However, our second approach to evaluation, including specific instructions (to evaluate the statement given as True or False), resulted in 100% concordance and specificity and a Cohen’s kappa of 1. This highlights the importance of prompts, which refer to appropriate instructions to interact with AI agents, that have been repeatedly noted since the release of ChatGPT to the public. They have become so relevant that practical guidelines have been developed specifically for healthcare professionals and their interaction with Conversational Agents [15]. This aspect must be emphasised when training users to implement LLMs in their medical practice.

We also noticed that, while conducting the second evaluation, if the model couldn’t access the bibliography because of a connection issue, it would still answer with its background knowledge, citing no references and likely using the default information from the pre-training received by GPT-4o. This would sometimes result in the resurfacing of mistakes, regardless of appropriate prompting. Therefore, users should monitor the models’ use of references, and when none are given, it is safer to repeat the process. Regenerating answers when they did not cite references during our experimentation corrected this mistake.

On the other hand, mistakes would also arise when the model was presented with many statements simultaneously, regardless of clear prompting. This is associated with the limit of information that the model can hold at once. The model performs a keyword search through the clinical sources provided and extracts information to answer the query; however, it cannot access and hold all the information simultaneously. Therefore, the model might answer incorrectly if the initial information extraction does not include the chunks (bits of information) necessary to retrieve a given statement. This highlights another critical avenue of user education: making queries one at a time to ensure optimal performance.

Nevertheless, analysis of results also showed that using specific prompting improves performance in retrieval completeness and usefulness without altering clinical understanding. A rare finding was that this prompting seemed to cause clinical reasoning to decrease. It is important to consider that retrieval is negatively affected when the model fails to reference the information. It occurs when there is a connection issue before the model can finish searching the bibliography. As stated, these cases can be handled by regenerating the response. Additionally, the issue of usefulness can be related to the ambiguity of some answers. This general manner of answering is not necessarily unfavourable but strongly emphasises the importance of clinician criteria when using conversational agents.

Regarding the four incorrect statements, the first two were about dysphagia in dementias and its management in Alzheimer’s patients. We realized that the assistant understood the clinical meaning of the word but did not have a functional understanding of it when associated with an altered state of mind in a statement but given no further instruction (straightforward approach). We realized that dysphagia was mentioned in less than 50% of the clinical guidelines used as a knowledge base for the AC. This represents a bias in the selection of information that results in a response that could potentially induce a harmful practice for the integrity and well-being of the patient. We expect to broaden the scope of guidelines included to relevant papers that could cover as much information as possible in the improved versions of this Conversational Assistant, outlined in the next paragraph.

The human evaluation revealed further insights into model performance and avenues for improvement. Although the model excels at the clinical understanding of the question and response completeness, it could improve retrieval and perceived usefulness of the answer. The improvement of the model is an ongoing process and includes testing it with laypeople (in this paper, the evaluators were not experts in medicine or Alzheimers) and the upcoming validation, which has two stages. The first one, where predefined questions will be assigned to general physicians (the CA’s target population) for them to interact with the Assistant, to be compared with interactions of a control group of physicians that will answer the same questions using more traditional internet resources, but excluding any another form of artificial intelligence, and the second one, where one group of neurologists specialized in dementia will evaluate the comparison of answers from the previous step, and a second group will evaluate the Assistant answers for the clinical reasoning, language, response appropriateness and potential to cause harm.

The application of LLMs to medical questions has been under investigation with closed and open source models like GPT-3.5, Llama 2, and MedPalm, obtaining passing scores on popular medical benchmarks. One study evaluated GPT-3.5, InstructGPT, Codez, Llama 2, Vicuna, Guanaco, Falcon, MPT and GPT-NeoX on medical board exam question datasets: the US Medical Licensing Examination (USMLE), which includes complex, real-world medical questions for medical professionals, and has a passing score of 60; the MedMCQA dataset, which is comprised of medical school entrance exams (passing score: 50) and PubMedQA (Human expert score, passing score 78) that is designed to test medical reading comprehension using PubMed abstracts annotated by experts. Models achieved scores of 50.3 to 86.5 in USMLE, 52.9 to 73.7 in MedMCQA and 77.4 to 81.2 in PubMedQA, reaching and surpassing the standard tests’ passing scores. They were also evaluated in terms of their capabilities to carry out correct reasoning steps, with a 70% success, correctly recall knowledge (72% success), and adequate reading comprehension (90% success) [16].

Studies have explored the specialisation of language models to medical areas using specific knowledge bases and testing on medical QA tasks. LLM specialisation has been explored in general medicine and tested against standard medical question-answering (QA) datasets from US Medical Board, MedQA-USMLE and MedMCQA. In this study, models that used medical textbooks as a knowledge base were compared to models using closed-book models, which are explicitly pre-trained for the medical domains and rely only on their internal knowledge, and Wikipedia-augmented models, that use Wikipedia knowledge to assist in the QA task in the MedQA-USMLE and MedMCQA. Llama, GPT-3.5 and GPT-4 obtained in the MedQA-USMLE 31.4, 51.3 and 81.7 respectively as closed-book models, 39.9, 54.2 and 81.5 as Wikipedia-Augmented Models and 42.2, 67.9 and 88.1 as Textbook-Augmented models, this trend was maintained for MedMCQA. Showcasing the potential of textbook-specific and enriching models beyond their internal knowledge [17].

Model specialization has also been explored for specific medical domains. For example, LiVersa for liver disease used retrieval-augmented generation (RAG) on 30 publicly available American Association for the Study of Liver Diseases guidelines and guidance documents. The model obtained a perfect score in a 10-question “yes” or “no” previously published knowledge study. It committed mistakes in justifications and rationales on three questions, highlighting its knowledge deficiencies due to limitations in the number and type of documents that can be used for RAG [18]. Another study developed Uro_Chat based on the European Association Guidelines using LlamaIndex for urology-oncology. The model is tested on 15 questions related to uro-oncology detailed in a previous publication and 5 specific questions from recent literature. Its performance is compared to ChatGPT-3.5 and ChatGPT-4. Uro_Chat responds adequately to all 15 questions and correctly addresses all five extra questions, while ChatGPT 3.5 and 4, despite providing well-written, plausible answers, may contain fundamentally wrong or inadequate information [19]. Similarly, our DACA is a specialised LLM that has demonstrated promising retrieval capability to build accurate answers to clinical questions, when prompted correctly, regarding Alzheimer’s and Dementia.

The improvement in CA responses, when given proper prompting, underlines the importance of user training and reiterates the use of DACA as a support tool in conjunction with human medical criteria. The data collected does not allow for a detailed analysis of the knowledge base, and questions remain on how the model uses this information, whether prioritizing some information over another or accessing more frequent guides in Spanish or English or representatives from specific regions. The use of three different knowledge scales aims to assess the model’s knowledge on many different avenues however, this doesn’t completely confirm if the knowledge base is a sufficiently representative sample of Alzheimer’s literature.

Additionally, this initial validation has yet to include clinicians. A deeper evaluation will be conducted with more complex questions and direct user feedback on the assistant’s ease of use, quality and utility. Furthermore, the assistant is still susceptible to making mistakes, especially if it loses connection before retrieval is completed. Handling these situations requires user criteria to resubmit the question.

## CONCLUSION

Despite the mentioned limitations, this study demonstrates the potential of DACA and specialised LLMs in the medical field as a firm step towards personalised medical care and up-to-date clinical information, particularly when medical knowledge is becoming increasingly vast and ever-changing.

## Data Availability

All data produced in the present study are available upon reasonable request to the authors

## Contributions

NCV: Validation methodology and execution. Editor, Research and manuscript writing

IL: Validation methodology and execution. Research and manuscript writing

MCV: Methodology execution, including information processing, AI design, experimentation, model output, results and interpretation

JM: Supervision of information of data processing, AI design and training and model output

JZ: Project management and supervision

TU: Expert revisor

AMB: Expert revisor

CB: Expert revisor

NM: Expert revisor

